# COVID-19 Vaccine Perceptions and Uptake in a National Prospective Cohort of Essential Workers

**DOI:** 10.1101/2021.10.20.21265288

**Authors:** Karen Lutrick, Holly Groom, Ashley L. Fowlkes, Kimberly Groover, Manjusha Gaglani, Patrick Rivers, Allison L. Naleway, Kimberly Nguyen, Meghan Herring, Kayan Dunnigan, Andrew Phillips, Joel Parker, Julie Mayo Lamberte, Khaila Prather, Matthew S. Thiese, Zoe Baccam, Harmony Tyner, Sarang Yoon

## Abstract

**Introduction:** In a multi-center prospective cohort of essential workers, we assessed knowledge, attitudes, and practices (KAP) by vaccine intention, prior SARS-CoV-2 positivity, and occupation, and their impact on vaccine uptake over time.

**Methods:** Initiated in July 2020, HEROES-RECOVER cohort provided socio-demographics and COVID-19 vaccination data. Using follow-up two surveys approximately three months apart, COVID-19 vaccine KAP, intention, and receipt was collected; the first survey categorized participants as reluctant, reachable, or endorsers.

**Results:** A total of 4,803 participants were included in the analysis. Most (70%) were vaccine endorsers, 16% were reachable, and 14% were reluctant. By May 2021, 77% had received at least one vaccine dose. KAP responses strongly predicted vaccine uptake, particularly positive attitudes about safety (aOR=5.46, 95% CI: 1.4-20.8) and effectiveness (aOR=5.0, 95% CI: 1.3-19.1). Participants prior SARS-CoV-2 infection were 22% less likely to believe the COVID-19 vaccine was effective compared with uninfected participants (aOR 0.78, 95% CI: 0.64-0.96). This was even more pronounced in first responders compared with other occupations, with first responders 42% less likely to believe in COVID-19 vaccine effectiveness (aOR=0.58, 95% CI 0.40-0.84). KAP responses shifted positively, with reluctant and reachable participant scores modestly increasing in positive responses for perceived vaccine effectiveness (7% and 12%, respectively) on the second follow-up survey; 25% of initially reluctant participants received the COVID-19 vaccine.

**Discussion:** Our study demonstrates attitudes associated with COVID-19 vaccine uptake and a positive shift in attitudes over time. First responders, despite potential high exposure to SARS-CoV-2, and participants with a history of SARS-CoV-2 infection were more vaccine reluctant.

**Conclusions:** COVID-19 vaccine KAP responses predicted vaccine uptake and associated attitudes improved over time. Perceptions of the COVID-19 vaccine can shift over time. Targeting messages about the vaccine’s safety and effectiveness in reducing SARS-CoV-2 virus infection and illness severity may increase vaccine uptake for reluctant and reachable participants.

## INTRODUCTION

The SARS-CoV-2 pandemic has resulted in high levels of morbidity and mortality in the US.^1^ In response, a global effort to develop COVID-19 vaccines generated evidence leading to the U.S. Food and Drug Administration (FDA) authorizing COVID-19 vaccines under an Emergency Use Authorization (EUA) mechanism, beginning in mid-December 2020.^2^ Essential workers, including healthcare personnel (HCP), first responders, and other frontline workers (FW), may be at an increased risk of SARS-CoV-2 infection because of their high rates of contact with patients, coworkers, or the general public^3-7^ and were prioritized to receive COVID-19 vaccines by the Centers for Disease Control and Prevention (CDC) Advisory Committee on Immunization Practices during initial, staggered distribution.

The COVID-19 vaccines have been shown to be safe and effective in adults and children ages 12 and older, but the initial high demand for vaccination has decreased.^8^ Prior to COVID-19 vaccine authorization and availability in December 2020, early studies in the United States (US) reported rates of willingness to receive the COVID-19 vaccine ranging widely from 40% to 75%.^9-18^ Additionally, first responders and FW have reported lower rates of vaccine acceptance than HCP.^12,14^ Common reasons for vaccine hesitancy included the novelty of the COVID-19 vaccines, concerns about potential adverse effects, and/or a distrust in government.^9-14^

There is some indication that COVID-19 vaccine acceptance has changed over time in cross-sectional surveys. ^12,19^ It is unclear how individual vaccination intention has evolved as the public, has gained more information regarding symptoms and outcomes of COVID-19 disease and risks and benefits of vaccinations.

Knowledge, attitudes, and practices (KAP) toward vaccination are often examined to understand factors associated with the acceptability of vaccines and inform strategies for increasing vaccine uptake.^20^ We have addressed these knowledge gaps with a multi-center prospective cohort of essential workers with the following objectives: 1) assess differences in KAP by vaccine intention, prior SARS-CoV-2 positivity, and occupation group; 2) examine KAP as predictors of vaccine uptake; and 3) assess individual-level change in KAP over time.

## METHODS

### Study Design & Population

The HEROES-RECOVER studies represent a national network of prospective cohorts, including Arizona Healthcare, Emergency Response and Other Essential Workers Surveillance Study (HEROES) and Research on the Epidemiology of SARS-CoV-2 in Essential Response Personnel (RECOVER) funded by the CDC with sites in Phoenix, Tucson, and other areas in Arizona; Miami, Florida; Duluth, Minnesota; Portland, Oregon; Temple, Texas; and Salt Lake City, Utah. Details of the protocols of the studies have been previously published.^21^ Ongoing enrollment began in July 2020 and included HCP, first responders, and FW who worked at least 20 hours per week and had routine occupational exposure to coworkers or the public.

Participants completed detailed epidemiologic surveys at enrollment and at approximately three-month intervals (Follow-up surveys 1 and 2); text message-based surveys were completed weekly and monitored illness or potential COVID-19 contact in the past 7 days. The study is ongoing, but for this analysis participants actively enrolled during the Follow-up 1 survey distribution were included, with their prior SARS-CoV-2 infection, COVID-19 vaccination, and KAP data through May 19, 2021 utilized for analysis.

To identify SARS-CoV-2 infections, participants self-collected mid-turbinate nasal swabs weekly for SARS-CoV-2 RT-PCR testing and provided blood specimens at enrollment and every subsequent three months (supplemental Figure 1 for study timing). Beginning in December 2020, participants were prompted to report uptake of COVID-19 vaccine; vaccine survey distribution was based on vaccine availability data from state and county health departments. Vaccination was verified by participant-provided vaccine cards, electronic medical records, or State Immunization Information Systems. All protocols were reviewed and approved by each site’s Institutional Review Boards; study participants provided informed consent for all study activities.

### Primary Outcomes

Vaccine intention and KAP questions were in two follow-up surveys: Follow-up survey 1 was distributed from late December 2020-February 2021, and Follow-up survey 2 -- from March 2021-May 2021. New enrollees during each follow-up period received the KAP questions at the time of enrollment.

Vaccine intention was derived using participants’ first response to the question, “What are the chances that you will get a COVID-19 vaccination?” and vaccination status at the time of Follow-up survey 1. Participants were grouped into three intention categories: 1) reluctant as those who answered, “almost zero chance”, or “very small chance”, and were unvaccinated, 2) reachable as those who answered “small chance”, “do not know”, or “moderate” and were unvaccinated, or 3) endorser as those who answered, “large chance”, “very large chance”, or “almost certain”, or were vaccinated at Follow-up survey 1. New vaccine receipt after Follow-up survey 1 was monitored and the participants’ vaccine intention group did not change based upon Follow-up survey 2 KAP responses.

Participants were asked six questions to assess the KAP constructs regarding COVID-19: knowledge of SARS-CoV-2 and COVID-19 vaccines; attitudes about safety, effectiveness, trust in the government, and perceived risk of becoming ill if they were not vaccinated (Table *1*Table *1*). Responses to each question were rated on a 5- to 7-level Likert scale indicating lowest to highest ranking.

**Table 1.** Knowledge, Attitude, and Practice (KAP) Questions.

| Topic | Question Text | Range |
| --- | --- | --- |
| **Vaccine Intention** | What are the chances that you will get a COVID-19 vaccination? | 8-point Likert (1=Don't know, 8=Almost certain) |
| **Chance of getting sick if not vaccinated** | If you are unable to or don't get a COVID-19 vaccination, what do you think your chance of getting sick with COVID-19 this year will be? | 7-point Likert (1=Almost zero, 8=Almost certain) |
| **Virus Knowledge** | How much do you know about the SARS-CoV-2 (COVID-19) virus and the illness it causes? | 5-point Likert (1=Nothing at all, 5=A great deal) |
| **Vaccine Knowledge** | How much do you know about the COVID-19 vaccine? Would you say...? | 5-point Likert (1=Nothing at all, 5=A great deal) |
| **Vaccine Safety** | How safe do you think the COVID-19 vaccine is? | 5-point Likert (1=Not at all, 5=Extremely safe) |
| **Vaccine Effectiveness** | How effective do you think the COVID-19 vaccine is in preventing you from getting sick with COVID-19? | 5-point Likert (1=Not at all, 5=Extremely effective) |
| **Trust in government** | I trust what the government says about the COVID-19 vaccine | 5-point Likert (1=Strongly disagree, 5=Strongly agree) |

### Predictors and Confounders

For models examining KAP differences and predictors of vaccination, socio-demographics, including gender, age, race, ethnicity, education, household income, occupation and occupational setting, and participant health status, including SARS-CoV-2 infection status, COVID-19 vaccination status, and medical history were included. HCP occupation categories are categorized as any individual that works in a hospital as “HCP inpatient”, any individual that works in any outpatient healthcare facility or long-term care facility as “HCP other”. We created two first responder categories: 1) firefighter (firefighters/EMS) and 2) other first responders (law enforcement, correctional officers, and border patrol). FW public-facing included individuals that work in education settings, retail, food service, and hospitality. Other FW include individuals that work in infrastructure, manufacturing, warehouse, utility, and transportation.

In models examining Objectives 1 and 2, COVID-19 contact data were reported as the number of hours spent at work (1) in any setting and in direct contact with individuals with suspected or confirmed COVID-19 and (2) the general public in the past 7 days. They also indicated the percent of time protective equipment (PPE) was used during this contact.

Participants were categorized as having had a SARS-CoV-2 infection prior to Follow-up survey 1 if they reported detection by antibody, antigen, or RT-PCR assay prior to enrollment, or if SARS-CoV-2 was detected by RT-PCR or an antibody test during the study.

For Objectives 2 and 3 (KAP change over time), KAP responses (defined above) were used as the primary predictors of interest.

### Statistical Analysis

We included all participants who completed the Follow-up 1 survey. Continuous measurements were expressed as means and standard deviations or median and interquartile range, as appropriate. Counts and percentages were used for categorical variables. Likert scores were dichotomized for each KAP question, using responses greater than midpoint as positive associations and midpoint and lower than the midpoint as neutral/negative associations (Table *1*).

We stratified socio-demographics, occupation and occupational setting, previous positivity, KAP responses by vaccine intention, and utilized chi-squared tests or one-way ANOVA tests to examine family-wise differences between the vaccine intention groups, with statistical significance based on p-values <0.05.

To examine KAP differences (Objective 1), we used unadjusted ordinal logistic regression to examine the relationship between each KAP question in the Follow-up 1 survey and vaccine intention, each occupation, and prior positivity. Bonferroni corrections adjusted for multiple comparisons and statistical significance based on 95% confidence intervals. We also used a difference in proportion test to test pair-wise differences in answers to KAP questions.

For KAP predictors (Objective 2), we utilized adjusted ordinal logistic regression to test the effect of each KAP on vaccine uptake when including socio-demographics, occupation and occupational setting, vaccine intention, and prior positivity together. Bonferroni corrections adjusted for the multiple comparisons and statistical significance based on 95% confidence intervals.

For KAP change (Objective 3), we tested differences in answers to KAP questions in Follow-up 2 compared to Follow-up 1 on a subset that completed both surveys. Chi-squared tests were used to determine statistically significant differences in each KAP question at Follow-up 2 compared to Follow-up 1, with significance based on p-values <0.05. We descriptively examined vaccine uptake and KAPs over time for each vaccine intention group. All statistical analyses were completed using R (version 4.0.4; R Foundation for Statistical Computing) and SAS (version 9.4; SAS Institute).

## RESULTS

### Overall Participants

December 2020 -February 2021, 4,803 (87%) of 5,527 participants responded to Follow-up survey 1; 1,105 (23%) HCP inpatient, 1,323 (28%) other HCP, 729 (15%) first responder firefighter, 255 (5%) other first responders, 990 (21%) FW Public, and 285 (6%) other FW (Table ***2***). Most participants were female (62%) and aged <45 years (58%). Additionally, 72% were non-Hispanic White, 14% Hispanic, 9% other, 3% Asian American/Pacific Islander, and 2% African American. Participants were highly educated, including 76% with at least a college degree, and only 15% percent reporting annual income less than $50,000. Participants were healthy, with only 24% reporting an underlying condition, most commonly hypertension (12%), asthma (9%), and diabetes (3%). At the time of the Follow-up 1 survey, 960 (20%) of participants had previously been infected with SARS-CoV-2. Total positive rates amongst FW and HCP were similar (25% and 22% respectively), with higher rates amongst first responders (32%). Thirty-six percent had received a COVID-19 vaccination at the time of the Follow-up 1 survey.

**Table 2.**
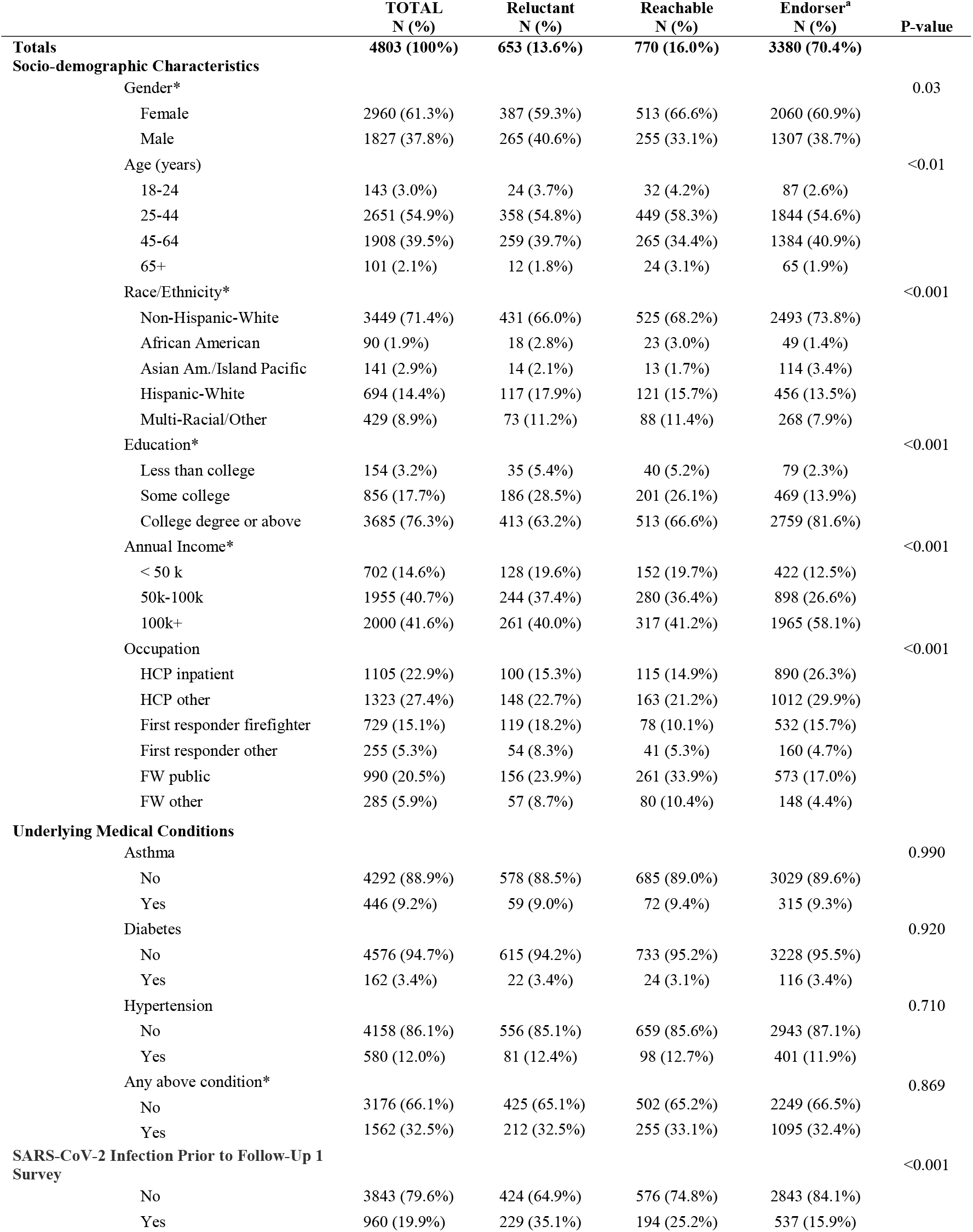
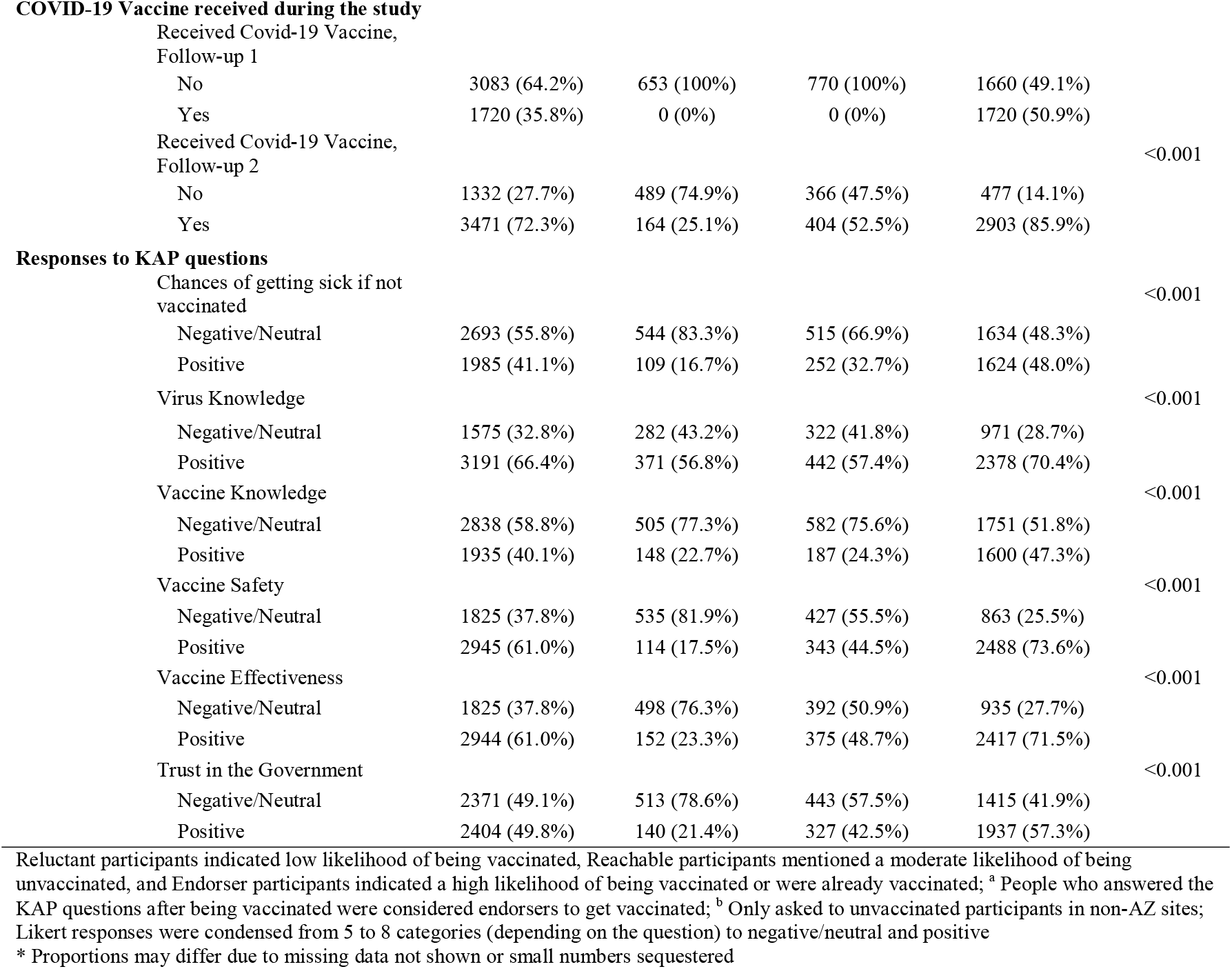
Descriptive Statistics, Stratified by Vaccine Intent Group in a Survey of Essential Workers December 2020 through May 2021

### Vaccination Intent

Most participants were categorized as endorsers (70%), having either indicated a high likelihood of intent to receive the COVID-19 vaccine (35%) or having already received it at the time of Follow-up 1 survey (36%); 16% of participants were considered reachable, and 14% reluctant. Prior SARS-CoV-2 infection was more common among reluctant (35%) and reachable participants (25%) compared with endorsers (16%). By May 19, 2021, 72% of participants hae received at least one dose of a COVID-19 vaccine (Table ***2***). Vaccine uptake varied by intention group, including reported COVID-19 vaccine receipt among 86% of endorsers, 53% of reachable, and 25% of reluctant.

#### Objective 1: KAP as predictor for vaccine uptake

After adjusting for socio-demographic factors, health status, and hours of direct contact with the public, KAP responses strongly predicted vaccine uptake. Participants reporting more positive attitudes about COVID-19 vaccine safety were 5.5 times more likely to receive a COVID-19 vaccine compared with those reporting more negative attitudes (aOR=5.46, 95% CI: 1.43-20.82) and 5 times as likely to receive a COVID-19 vaccine among participants reporting a belief that the vaccine is effective (aOR=4.98 95% CI: 1.30-19.14) (Table 3).

**Table 3.** Difference in Knowledge, Attitude, and Practice (KAP) Questions Stratified by Vaccination Status, Intention Group, Occupation, and Prior SARS-CoV-2 Positivity in a Cohort of Essential Workers (N=4803)^a^

|  | Unadjusted |  | Adjusted<sup>b</sup> |  |
| --- | --- | --- | --- | --- |
|  | OR | 95% CI | OR | 95% CI |
| **Vaccinated during the study (not vaccinated is the reference group)** |  |  |  |  |
| Virus Knowledge | 1.58 | 1.40 – 1.79 |  |  |
| Vaccine Knowledge | 2.49 | 2.17 – 2.87 |  |  |
| Vaccine Safety | 9.81 | 8.42 – 11.44 | 5.46 | 1.43 – 20.82 |
| Vaccine Effectiveness | 8.29 | 7.10 – 9.67 | 4.98 | 1.30 – 19.14 |
| Trust in government | 4.40 | 3.87 – 5.00 |  |  |
| Chances of getting sick | 4.15 | 3.58 – 4.81 |  |  |
| **By Intention Group (Endorser is the reference Group)** |  |  |  |  |
| Reluctant |  |  |  |  |
| Virus Knowledge | 0.53 | 0.45 – 0.62 |  |  |
| Vaccine Knowledge | 0.30 | 0.26 – 0.35 | 0.49 | 0.34 – 0.72 |
| Vaccine Safety | 0.08 | 0.06 – 0.09 | 0.23 | 0.15 – 0.33 |
| Vaccine Effectiveness | 0.12 | 0.10 – 0.14 | 0.32 | 0.22 – 0.48 |
| Trust in government | 0.20 | 0.17 – 0.23 | 0.43 | 0.30 – 0.61 |
| Chances of getting sick | 0.23 | 0.20 – 0.27 | 0.48 | 0.32 – 0.74 |
| Reachable |  |  |  |  |
| Virus Knowledge | 0.52 | 0.45 – 0.60 |  |  |
| Vaccine Knowledge | 0.34 | 0.30 – 0.40 | 0.53 | 0.30 – 0.96 |
| Vaccine Safety | 0.33 | 0.28 – 0.38 | 0.56 | 0.31 – 1.00 |
| Vaccine Effectiveness | 0.40 | 0.35 – 0.47 |  |  |
| Trust in government | 0.58 | 0.51 – 0.67 |  |  |
| Chances of getting sick | 0.59 | 0.51 – 0.68 |  |  |
| **Occupation (HCP inpatient is the reference group)** |  |  |  |  |
| HCP other |  |  |  |  |
| Virus Knowledge | 0.81 | 0.70 – 0.94 |  |  |
| Vaccine Knowledge | 0.97 | 0.84 – 1.12 |  |  |
| Vaccine Safety | 0.91 | 0.79 – 1.06 |  |  |
| Vaccine Effectiveness | 1.02 | 0.87 – 1.18 |  |  |
| Trust in government | 0.98 | 0.85 – 1.13 |  |  |
| Chances of getting sick | 0.89 | 0.77 – 1.03 |  |  |
| First responder firefighter |  |  |  |  |
| Virus Knowledge | 0.37 | 0.31 – 0.44 | 0.48 | 0.39 – 0.59 |
| Vaccine Knowledge | 0.43 | 0.36 – 0.51 | 0.57 | 0.46 – 0.71 |
| Vaccine Safety | 0.43 | 0.36 – 0.51 | 0.60 | 0.48 – 0.74 |
| Vaccine Effectiveness | 0.41 | 0.34 – 0.49 | 0.61 | 0.49 – 0.76 |
| Trust in government | 0.62 | 0.52 – 0.73 |  |  |
| Chances of getting sick | 0.72 | 0.61 – 0.85 |  |  |
| First responder other |  |  |  |  |
| Virus Knowledge | 0.20 | 0.15 – 0.25 | 0.25 | 0.18 – 0.36 |
| Vaccine Knowledge | 0.19 | 0.15 – 0.25 | 0.34 | 0.24 – 0.49 |
| Vaccine Safety | 0.34 | 0.26 – 0.43 | 0.46 | 0.32 – 0.67 |
| Vaccine Effectiveness | 0.41 | 0.32 – 0.53 | 0.58 | 0.40 – 0.84 |
| Trust in government | 0.48 | 0.37 – 0.60 | 0.67 | 0.47 – 0.95 |
| Chances of getting sick | 0.71 | 0.56 – 0.91 |  |  |
| FW Public |  |  |  |  |
| Virus Knowledge | 0.30 | 0.26 – 0.36 | 0.41 | 0.34 – 0.50 |
| Vaccine Knowledge | 0.30 | 0.25 – 0.35 | 0.41 | 0.33 – 0.50 |
| Vaccine Safety | 0.65 | 0.55 – 0.76 |  |  |
| Vaccine Effectiveness | 0.75 | 0.64 – 0.88 | 1.25 | 1.02 – 1.53 |
| Trust in government | 0.95 | 0.82 – 1.11 | 1.38 | 1.14 – 1.68 |
| Chances of getting sick | 0.94 | 0.81 – 1.10 | 1.33 | 1.13 – 1.56 |
| FW other |  |  |  |  |
| Virus Knowledge | 0.28 | 0.22 – 0.35 | 0.41 | 0.35 – 0.49 |
| Vaccine Knowledge | 0.36 | 0.28 – 0.45 | 0.49 | 0.41 – 0.57 |
| Vaccine Safety | 0.59 | 0.47 – 0.75 |  |  |
| Vaccine Effectiveness | 0.72 | 0.56 – 0.91 | 1.49 | 1.26 – 1.77 |
| Trust in government | 0.86 | 0.68 – 1.08 |  |  |
| Chances of getting sick | 0.52 | 0.41 – 0.65 |  |  |
| **Prior SARS-CoV-2 Infection (No known prior infection as the reference group)** |  |  |  |  |
| Virus Knowledge | 0.91 | 0.85 – 0.98 |  |  |
| Vaccine Knowledge | 0.62 | 0.57 – 0.68 | 0.78 | 0.64 – 0.95 |
| Vaccine Safety | 0.51 | 0.47 – 0.55 |  |  |
| Vaccine Effectiveness | 0.48 | 0.44 – 0.52 | 0.78 | 0.64 – 0.96 |
| Trust in government | 0.62 | 0.58 – 0.67 |  |  |
| Chances of getting sick | 0.46 | 0.42 – 0.51 | 0.68 | 0.56 – 0.84 |

<sup>a</sup> P-values not reported due to inconsistencies that occur with multi-level categorical variables.
Statistical significance based on 95% confidence intervals.
<sup>b</sup> Non-significant adjusted point estimates and confidence intervals not reported. Bonferroni corrections were used for each of vaccination status, intention group, occupation, and prior positivity. The model was adjusted for socio-demographics, occupation and occupational setting, vaccine intention, and prior positivity for SARS-CoV-2 infection.

**Table 4.**
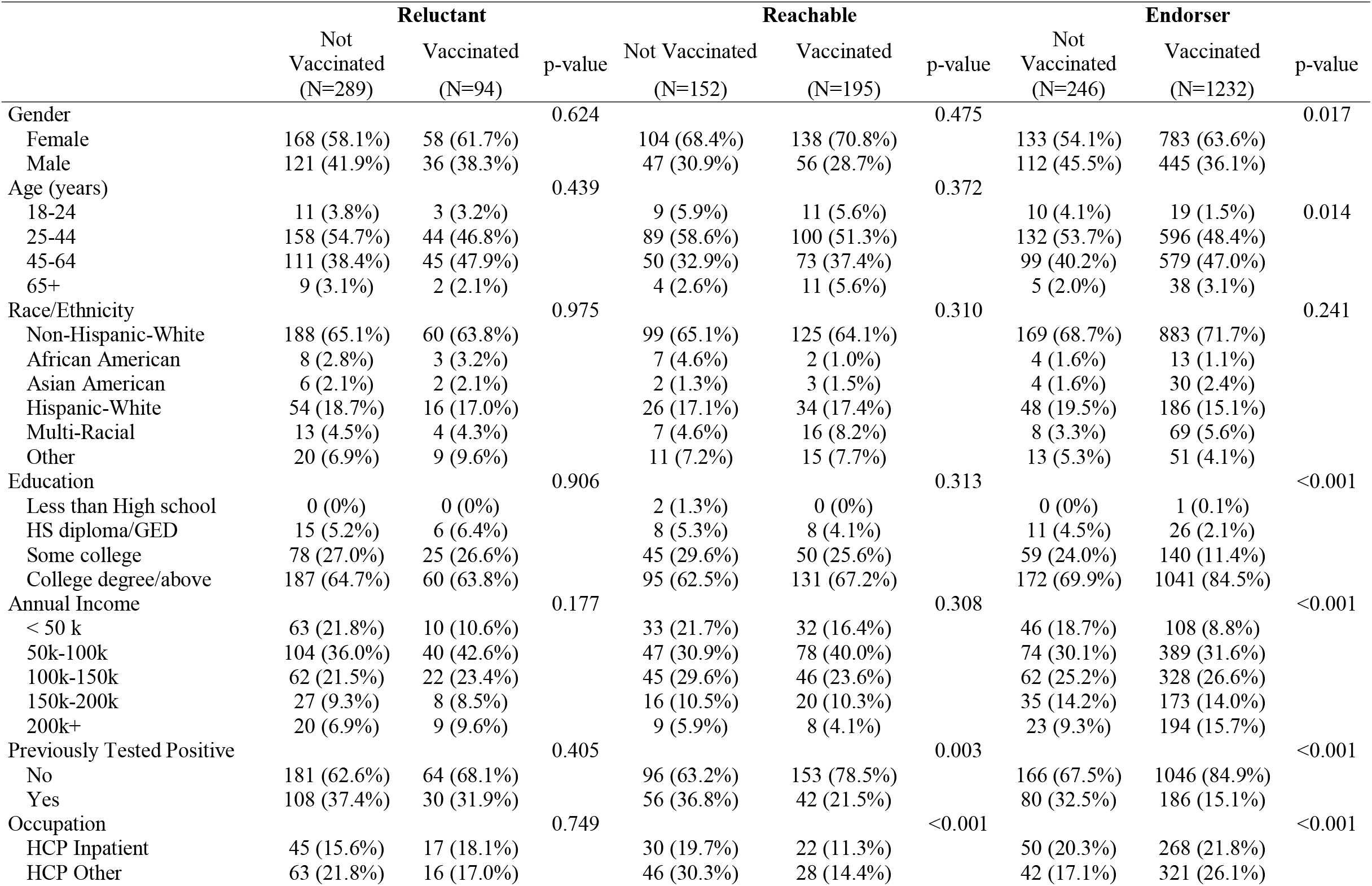
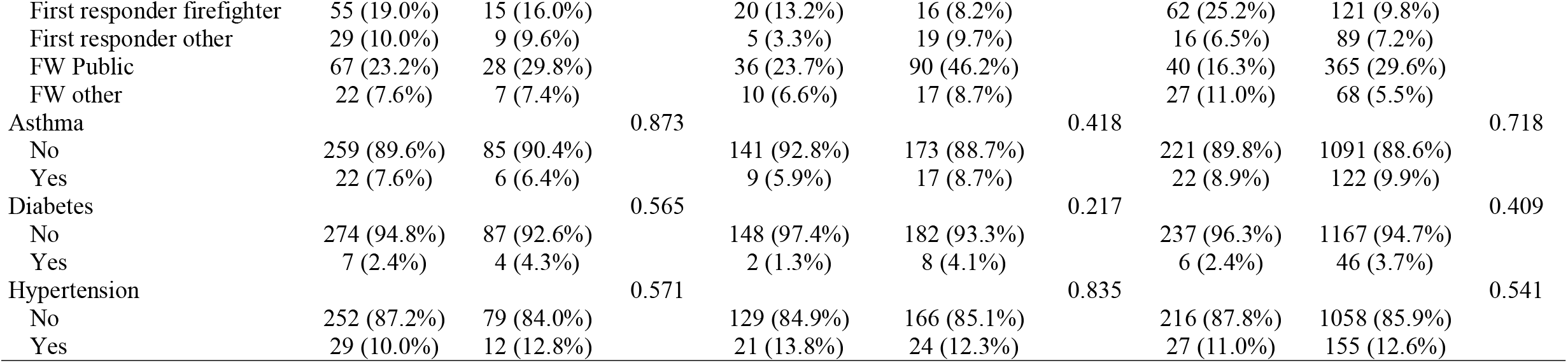
Demographics of Vaccine Intention Groups, Stratified by Vaccination Status at Time of Follow-up Survey 2 in a Cohort of Essential Workers

**Table 5.**
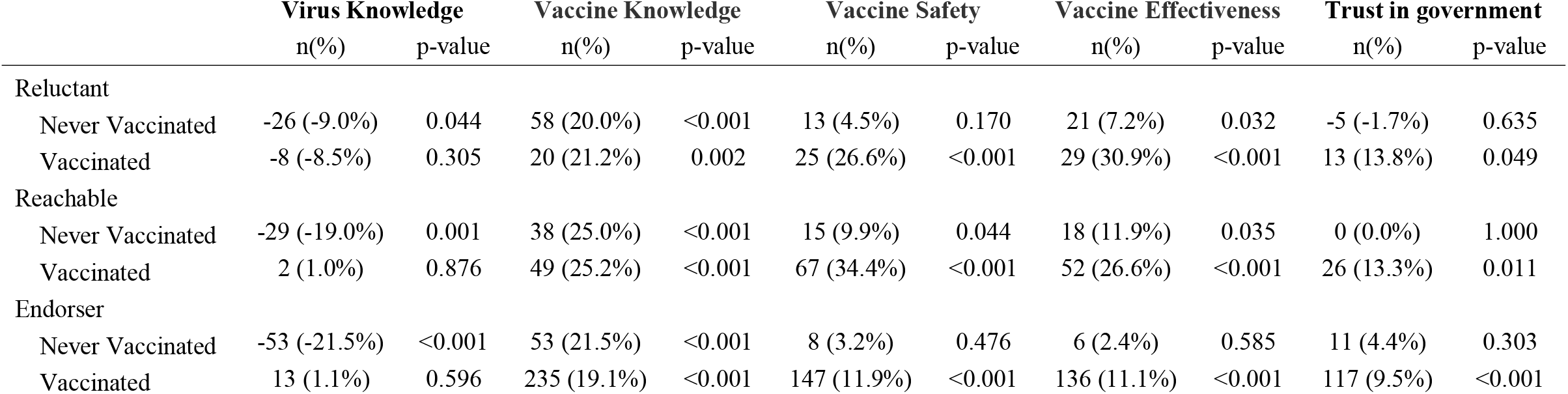
Change in Positive Response to Knowledge, Attitude, and Practice (KAP) Questions by Intention and Actual Vaccination from Follow-up Survey 1 to Follow-up Survey 2

#### Objective 2: KAP responses by intention group, prior SARS-CoV-2 infection and occupation

##### Vaccine Intention Groups

Only 17% of reluctant (n=109) and 33% of reachable participants (n=252) reported concern about getting sick if unvaccinated compared with 48% of endorsers (n=1624) (Table 2). Similarly, reluctant and reachable participants were more likely to report negative attitudes about vaccine safety (82% and 56%, respectively), vaccine effectiveness (76% and 51%, respectively), and trust in the government (79% and 58%, respectively).

Reachable participants were about half as likely and reluctant participants were substantially less likely to perceive the COVID-19 vaccines as safe compared to endorsers (aOR =0.56, 95% CI 0.31-1.00 and aOR=0.33, 95% CI: 0.28-0.38, respectively) (**Error! Reference source not found**.). Similarly, reluctant participants were 68% (aOR=0.32, 95% CI: 0.22-0.48) less likely than endorsers to perceive that the vaccine was effective. Interestingly, reachable participants were less likely to report knowledge about the COVID-19 vaccine than reluctant participants (aOR=0.53, 95% CI: 0.30-0.96 and aOR=0.49 95% CI: 0.34-0.75, respectively).

##### Prior SARS-CoV-2 Infection

Among 960 (20%) participants who reported SARS-CoV-2 infection prior to enrollment, 24% (n=229) were categorized as reluctant, 20% as reachable (n=194), and 56% (n=537) as endorsers (Table ***2***). COVID-19 vaccination through the study period was less common among participants with prior SARS-CoV-2 infection (n=576, 59%) compared to those without prior infection (n=3188, 82%). In the adjusted models, participants with prior SARS-CoV-2 infection were 32% less likely to be concerned about getting sick if not vaccinated (aOR 0.68, 95% CI: 0.56-0.84) and 22% less likely to believe the COVID-19 vaccine was effective (aOR 0.78, 95%CI: 0.64-0.96) compared with uninfected participants. Interestingly, there were no significant differences in perceived virus knowledge, vaccine safety, or trust in government by infection status in the adjusted models (Table 3).

##### Occupation

Overall, few HCP were COVID-19 vaccine reluctant, including 9% of HCP inpatient and 11% other HCP. Among first responders, subcategorization showed slight differences, with 16% of firefighters reluctant compared with 21% of other first responders. Similarly, 16% of public-facing FW and 20% of other FW were reluctant. Responses to KAP questions differed across occupations but were aligned with overall responses for vaccine intent groups that combine participants.

There was little difference between occupational subcategories of HCP or first responders in the adjusted models (Table 3**Error! Reference source not found**.). Firefighters and other first responder were each approximately 40% less likely than inpatient HCP to believe the COVID-19 vaccine was effective (aOR=0.58, 95% CI 0.40-0.84 and aOR=0.61, 95% CI 0.49-0.76, respectively). The other FW category was 51% more likely to believe the COVID-19 vaccine was effective compared to inpatient HCP (aOR=1.49, 95% CI 1.26-1.77), followed by public-facing FW (aOR=1.25, 95% CI 1.02-1.53) (**Error! Reference source not found**.).

#### Objective 3. KAP change over time

To evaluate change in KAP over time, 2017 (49%) participants that completed both Follow-up 1 and 2 surveys were included. Among initially 383 reluctant participants, 94 (25%) received COVID-19 vaccine; 195 (56%) reachable and 1,232 (83%) endorsers were also vaccinated. Demographic characteristics among reluctant and reachable participants who were vaccinated after initial categorization did not differ from unvaccinated participants. Among endorsers, unvaccinated participants were more likely to be male (p=0.017), younger (p=0.014)), and firefighters (p<0.001) than endorsers that were vaccinated (Table).

When evaluating KAP over time, reluctant participants that did not get vaccinated had a 9% decrease in positive responses to questions about their knowledge of the virus between Follow-up survey 1 and 2 (**Error! Reference source not found**.). The change in positive response to vaccine knowledge increased by 20% and 21% for the non-vaccinated and vaccinated, respectively. However, there was a 7% increase in positive response toward vaccine effectiveness in those that were not vaccinated compared to a 31% increase for vaccinated participants.

Participants in the reachable and endorser vaccine intent groups also showed decreases in positive responses for knowledge about the virus between the two time points (−19% and −22%, respectively), with higher percentages of participants reporting negative/neutral responses during Follow-up Survey 2 (**Error! Reference source not found**.). The reachable group had large increases in positive responses for questions about vaccine knowledge (25% of vaccinated, 25% of unvaccinated), vaccine safety (10% of vaccinated, 34% of unvaccinated), and vaccine effectiveness (12% of vaccinated, 27% of unvaccinated).

## DISCUSSION

The HEROES-RECOVER prospective cohort provided a unique opportunity to examine COVID-19 vaccine knowledge, attitudes, and practices longitudinally in a large population of essential workers with high occupational COVID-19 exposure. The prospective design allowed for assessment of vaccination intent as well as vaccine uptake.

We found KAP responses were strongly associated with vaccine uptake. Our cohort largely reported more positive attitudes toward the COVID-19 vaccine than other national cohorts,^12-15^ with more than two-thirds of participants expressing strong intent to be vaccinated. We also found strong associations between KAP responses and vaccine intention groups, with vaccine reluctant participants more likely to have negative attitudes towards safety and effectiveness and less likely to be vaccinated. While we found that a substantial proportion of our high-risk cohort population reported an initial reluctance to receive COVID-19 vaccine, ultimately one quarter of those reluctant were vaccinated by May 19, 2021.

### Vaccine Reluctance

First responders and participants with prior SARS-CoV-2 infection were more likely to be reluctant to receive the COVID-19 vaccine than other groups. First responders had the highest percentage of vaccine reluctant participants, especially the non-firefighter subcategory. Among endorsers, other first responders also had the lowest vaccination rates. This hesitancy towards the vaccine was also represented in lower perceptions of vaccine safety, effectiveness, and trust in government.

Participants with prior SARS-CoV-2 infection were less likely to receive the COVID-19 vaccine and make up more than one-third of the vaccine reluctant group and one-quarter of the reachable group. It is not surprising that participants previously positive for COVID-19 are less concerned about getting sick again, but better understanding why they report fewer positive attitudes toward vaccine safety and effectiveness will be important in persuading them to get vaccinated.^22,23^ Additional studies highlighting the benefits of vaccination for those with prior infection, may help to stress the importance of vaccination among this group.^24^

These findings are consistent with other vaccine acceptability studies done nationally^14,19^ and suggest that these negative attitudes persisted even after more data became available on the safety and efficacy of available vaccines.

### KAPs and Vaccine Uptake

Across intent to vaccinate, demographics, occupation, and prior SARS-CoV-2 infection groups, three KAP domains were consistently correlated with intent to vaccinate and vaccine uptake: safety, effectiveness, and the chance of getting sick if not vaccinated. We found knowledge about the SARS-CoV-2 virus, or the COVID-19 vaccine had no association with vaccine uptake. It is difficult to ascertain whether participants who perceive themselves to be knowledgeable are truly informed, but attitudes about vaccine safety and effectiveness appear to be more informative of individual intentions to vaccinate. Vaccination efforts that highlight vaccine safety and effectiveness may have a stronger influence on vaccination uptake than general or historical information. We found positive attitudes align with vaccine uptake and imply that KAP assessments to gauge a population’s intentions or concerns in advance of vaccination campaigns is critical.

Unsurprisingly, the majority of HCP were endorsers of the vaccine, and the vast majority received the COVID-19 vaccine. Some HCP occupational groups have low vaccination coverage nationally,^17,18^ and so our study population may not be representative of those groups. Other frontline workers, which for this study included teachers, retail workers, and manufacturing were not as positive towards the vaccine as HCP, though the vast majority were still considered endorsers and reachable and were vaccinated at higher rates than first responders. The COVID-19 pandemic has clearly demonstrated the critical nature of the essential worker role and need for additional investigations.

### KAP Change Over Time

Utilizing the prospective cohort, we were able to examine shifts in KAP over time, subgrouping vaccinated versus unvaccinated participants. The KAP factors that were most connected to vaccination remained influential over time. We identified more positive shifts in responses between the two time points in those participants ultimately vaccinated, specifically in response to perceived safety and effectiveness across all intention groups. Interestingly, even those participants that were not vaccinated demonstrated a positive increase in perceived vaccine safety and effectiveness over the three-month period.

Our findings are consistent with other studies conducted prior to COVID-19 vaccine authorization and availability.^14,16^ While vaccine intent was assessed in our study after the FDA granted EUA, our findings capture an initial uncertainty that was seemingly overcome with time and positive findings for vaccine safety and effectiveness.^12^

### Limitations

This study is subject to several limitations. First, the follow-up surveys were spread out over about six weeks due to site’s individual IRB timelines. As the level of information available evolved quickly during the study period, participants at sites where the follow-up surveys were administered later may have had access to a meaningfully different amount, or quality, of information. Secondly, all KAPs are self-reported and there may be a disconnect between perceived knowledge and actual level of knowledge. Next, while we are confident KAPs are successfully captured in our cohorts at the time of administration, due to the novelty of the COVID-19 vaccine, KAPs will likely continue to change and evolve past this analysis period. Finally, the mechanism prompting change in KAPs is not captured, so it is difficult to know why certain KAPs changed as they did over time, e.g., the change in certain KAPs between the two follow-up surveys may have been due to increased numbers of participants receiving the vaccine with few documented serious adverse event rates, increased access to information and disease/vaccine literacy, changes in national and local COVID-19 incidence. The demographic characteristics of the group that answered Follow-up 2 different slightly from those that completed Follow-up 1: there were more female participants (64% vs 60%), they were older (45% 40-65 years of age compared to 36%), and there were higher percentages of FW (36% vs 20%) and lower percentages of HCP (44% vs 58%). Race/ethnicity, education, and income were similar between the two groups. We did not differentiate between individual COVID-19 vaccine products in this analysis.

## Data Availability

All data produced in the present study are available upon reasonable request to the authors

## PUBLIC HEALTH IMPLICATION

The HEROES-RECOVER cohort provides valuable insight into the perceptions and intentions of essential workers receiving the COVID-19 vaccine. With the current increase in cases, encouraging high-risk occupational groups to receive the COVID-19 vaccine is a critical next step. Our findings indicate that perceptions of the COVID-19 vaccine can shift over time and suggest that focusing on clear messages about the vaccine’s safety and effectiveness in reducing SARS-CoV-2 virus infection and illness severity may increase vaccine uptake for reluctant and reachable participants. Targeted messaging by key stakeholders and healthcare providers for participants with prior infection and in occupations with low vaccine coverage and low trust in the government (like first responders) would be especially useful.

## LIST OF ABBREVATIONS

FDA: U.S. Food and Drug Administration
CDC: Centers for Disease Control and Prevention
EUA: Emergency Use Authorization
KAP: Knowledge, attitudes, and practices
HEROES: Arizona Healthcare, Emergency Response and Other Essential Workers Surveillance
RECOVER: Study and Research on the Epidemiology of SARS-CoV-2 in Essential Response Personnel
H-R: HEROES-RECOVER
HCP: Health care personnel
FW: Frontline workers
PPE: Personal protective equipment

## Statement of Contributions

K Lutrick, H Groom, A Fowlkes, K Groover, P Rivers, K Nguyen, M Herring, J Mayo Lamberte, K Prather, and S Yoon conceptualized the study and drafted the manuscript with the help of Z Baccam. J Parker and P Rivers conducted the statistical analysis. M Gaglani, A Naleway, K Dunnigan, A Phillips, M Thiese, and H Tyner were responsible for review and revision of the manuscript. All authors read and approved of the final manuscript.

## Acknowledgements

Supported by the National Center for Immunization and Respiratory Diseases and the Centers for Disease Control and Prevention (contracts 75D30120R68013 to Marshfield Clinic Research Institute, 75D30120C08379 to the University of Arizona, and 75D30120C08150 to Abt Associates).

Mark G. Thompson, Lauren Grant, Young M. Yoo, Gregory Joseph, Josephine Mak, Monica Dickerson, Suxiang Tong, John Barnes, Eduardo Azziz-Baumgartner, Melissa L. Arvay, Preeta Kutty, Alicia M. Fry, Lenee Blanton, Jill Ferdinands, Anthony Fiore, Aron Hall, Adam MacNeil, L. Clifford McDonald, Mary Reynolds, Sue Reynolds, Stephanie Schrag, Nong Shang, Robert Slaughter, Matthew J. Stuckey, Natalie Thornburg, Jennifer Verani, Vic Veguilla, Rose Wang, Bao-Ping Zhu, William Brannen, Stephanie Bialek, CDC; Jefferey L. Burgess, Shawn Beitel, Patrick Rivers, Xiaoxiao Sun, Joe K. Gerald, Katherine Ellingson, Ed Bedrick, Janko Nikolich-Žugich, Genesis Barron, Dimaye Calvo, Esteban Cardona, Andrea Carmona, Alissa Coleman, Emily Cooksey, Kiara Earley, Natalie Giroux, Sofia Grijalva, Allan Guidos, Adrianna Hernandez, James Hollister, Theresa Hopkins, Rezwana Islam, Krystal Jovel, Olivia Kavanagh, Jonathan Leyva, Sally Littau, Amelia Lobos, James Lopez, Veronica Lugo, Jeremy Makar, Taylor Maldonado, Enrique Marquez, Allyson Munoz, Assumpta Nsengiyunva, Joel Parker, Jonathan Perez Leyva, Alexa Roy, Saskia Smidt, Isabella Terrazas, Tahlia Thompson, Heena Timsina, Erica Vanover, Mandie White, April Yingst, Kenneth Komatsu, Elizabeth Kim, Karla Ledezma, University of Arizona, Arizona Department of Health Services; David Engelthaler, Translational Genomics Research Institute; Lauren E.W. Olsho, Danielle R. Hunt, Laura J. Edwards, Meredith G. Wesley, Tyler C. Morrill, Brandon P. Poe, Brian Sokol, Andrea Bronaugh, Tana Brummer, Hala Deeb, Rebecca Devlin, Sauma Doka, Tara Earl, Jini Etolue, Deanna Fleary, Jessica Flores, Chris Flygare, Isaiah Gerber, Louise Hadden, Jenna Harder, Lindsay LeClair, Nancy McGarry, Peenaz Mistry, Steve Pickett, Khaila Prather, David Pulaski, Rajbansi Raorane, Meghan Shea, John Thacker, Matthew Trombley, Pearl Zheng, Chao Zhou, Abt Associates; Spencer Rose, Tnelda Zunie, Michael E. Smith, Kempapura Murthy, Nicole Calhoun, Claire Mathenge, Arundhati Rao, Manohar Mutnal, Linden Morales, Shelby Johnson, Alejandro Arroliga, Madhava Beeram, Joel Blais, Jason Ettlinger, Angela Kennedy, Natalie Settele, Rupande Patel, Elisa Priest, Jennifer Thomas, Baylor Scott & White Health; Jennifer L. Kuntz, Yolanda Prado, Daniel Sapp, Mi Lee, Chris Eddy, Matt Hornbrook, Danielle Millay, Dorothy Kurdyla, Ambrosia Bass, Kristi Bays, Kimberly Berame, Cathleen Bourdoin, Carlea Buslach, Jennifer Gluth, Kenni Graham, Tarika Holness Enedina Luis, Abreeanah Magdaleno, DeShaun Martin, Joyce Smith-McGee, Martha Perley, Sam Peterson, Aaron Piepert, Krystil Phillips, Joanna Price, Sperry Robinson, Katrina Schell, Emily Schield, Natosha Shirley, Anna Shivinsky, Britta Torgrimson-Ojerio, Brooke Wainwright, Shawn Westaway, Kaiser Permanente Northwest; Jennifer Meece, Elisha Stefanski, Lynn Ivacic, Jake Andreae, Adam Bissonnette, Krystal Boese, Michaela Braun, Cody DeHamer, Timothy Dziedzic, Joseph Eddy, Heather Edgren, Wayne Frome, Nolan Herman, Mitchell Hertel, Erin Higdon, Rosebud Johnson, Steve Kaiser, Tammy Koepel, Sarah Kohn, Taylor Kent, Thao Le, Carrie Marcis, Megan Maronde, Isaac McCready, Nidhi Mehta, Daniel Miesbauer, Anne Nikolai, Brooke Olson, Lisa Ott, Cory Pike, Nicole Price, Christopher Reardon, Logan Schafer, Rachel Schoone, Jaclyn Schneider, Tapan Sharma, Melissa Strupp, Janay Walters, Alyssa Weber, Reynor Wilhorn, Ryan Wright, Benjamin Zimmerman, Marshfield Clinic Research Laboratory; Angela Hunt, Jessica Lundgreen, Karley Respet, Jennifer Viergutz, Daniel Stafki, St. Luke’s Regional Health Care System; Alberto J. Caban-Martinez, Natasha Schaefer-Solle, Paola Louzado Feliciano, Carlos Silvera, Karla Montes, Cynthia Beaver, Katerina Santiago, University of Miami; Rachel T. Brown, Camie Schaefer, Arlyne Arteaga, Matthew Bruner, Daniel Dawson, Emilee Eden, Jenna Praggastis, Joseph Stanford, Jeanmarie Mayer, Marcus Stucki, Riley Campbell, Kathy Tran, Madeleine Smith, Braydon Black, Madison Tallman, Chapman Cox, Derrick Wong, Michael Langston, Adriele Fugal, Fiona Tsang, Maya Wheeler, Gretchen Maughan, Taryn Hunt-Smith, Nikki Gallacher, Anika DSouza, Trevor Stubbs, Iman Ibrahim, Ryder Jordin, University of Utah; Marilyn J. Odean, Whiteside Institute for Clinical Research; Allen Bateman, Erik Reisdorf, Kyley Guenther, Erika Hanson, Wisconsin State Laboratory of Hygiene; the HEROES-RECOVER participants.

## Supplemental Appendix

**Supplemental Figure 1.**
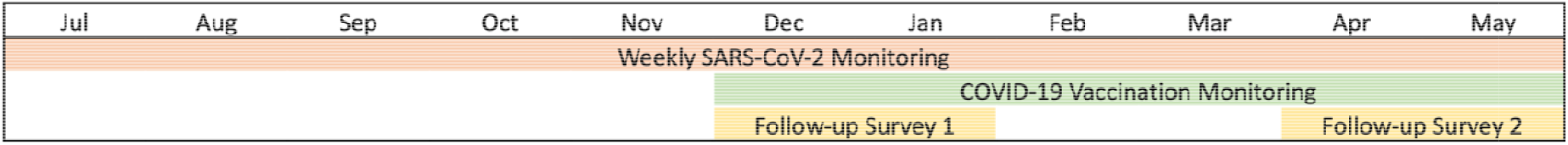
Timeline of key HEROES and RECOVER study activities July 2020-May 2021.

**Supplemental Figure 2.**
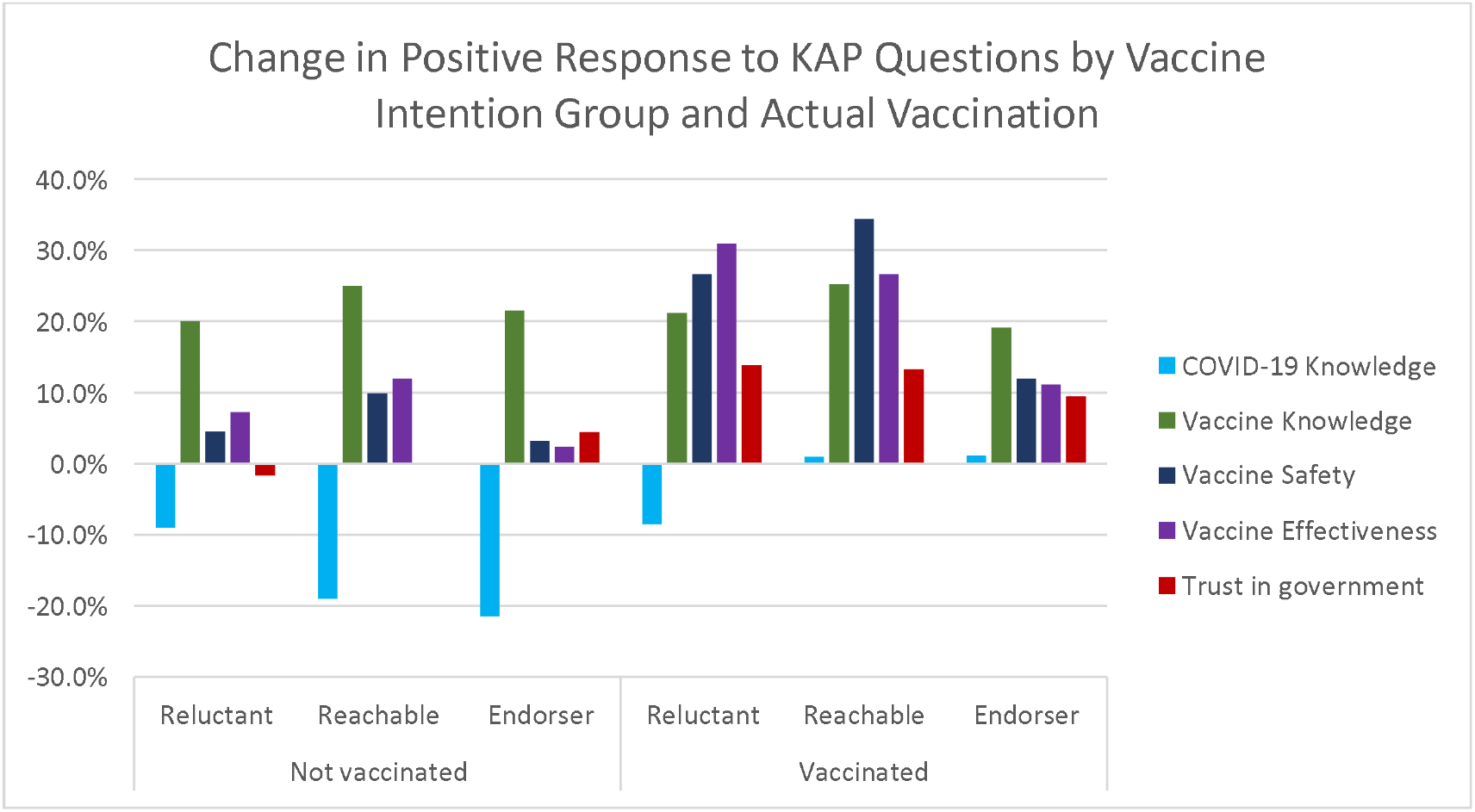
Change in Positive Response to Knowledge, Attitude, and Practice (KAP) Questions from Follow-Up Survey 1 to Follow-Up Survey 2, by Intention and Actual Vaccination.

**Supplemental Table 1.** KAP by Intention group, Occupation, and prior SARS-CoV-2 Infection.

|  | **Chance of**<br>**getting sick if**<br>**not vaccinated** | **Virus**<br>**Knowledge** | **Vaccine**<br>**Knowledge** | **Vaccine**<br>**Safety** | **Vaccine**<br>**Effectiveness** | **Trust in**<br>**government** |
| --- | --- | --- | --- | --- | --- | --- |
|  | Mean (SD) | Mean (SD) | Mean (SD) | Mean (SD) | Mean (SD) | Mean (SD) |
| **COVID-19 Vaccine Intention** |  |  |  |  |  |  |
| Reluctant | 3.47 (1.40) | 3.66 (0.876) | 2.97 (0.919) | 2.83 (0.846) | 2.98 (0.849) | 2.56 (1.19) |
| Reachable | 4.23 (1.37) | 3.64 (0.851) | 2.98 (0.843) | 3.50 (0.805) | 3.54 (0.774) | 3.32 (1.14) |
| Endorser | 4.62 (1.47) | 3.95 (0.839) | 3.57 (0.886) | 3.93 (0.794) | 3.88 (0.775) | 3.64 (1.20) |
| **Occupation** |  |  |  |  |  |  |
| HCP inpatient | 4.54 (1.48) | 4.14 (0.766) | 3.62 (0.882) | 3.87 (0.838) | 3.83 (0.798) | 3.54 (1.22) |
| HCP other | 4.44 (1.47) | 4.04 (0.805) | 3.60 (0.898) | 3.82 (0.884) | 3.82 (0.868) | 3.52 (1.26) |
| First responder firefighter | 4.26 (1.52) | 3.71 (0.833) | 3.20 (0.896) | 3.48 (0.907) | 3.43 (0.843) | 3.20 (1.24) |
| First responder other | 4.26 (1.52) | 3.40 (0.873) | 2.84 (0.869) | 3.35 (0.913) | 3.43 (0.860) | 3.02 (1.24) |
| FW public | 4.49 (1.51) | 3.61 (0.861) | 3.04 (0.859) | 3.68 (0.875) | 3.70 (0.812) | 3.50 (1.22) |
| FW other | 4.00 (1.46) | 3.57 (0.868) | 3.11 (0.840) | 3.66 (0.845) | 3.70 (0.800) | 3.43 (1.25) |
| **Prior SARS-CoV-2 Infection** |  |  |  |  |  |  |
| No | 4.52 (1.43) | 3.87 (0.848) | 3.40 (0.912) | 3.78 (0.867) | 3.77 (0.813) | 3.51 (1.23) |
| Yes | 3.91 (1.67) | 3.82 (0.885) | 3.17 (0.935) | 3.46 (0.916) | 3.44 (0.897) | 3.18 (1.25) |

*All relationships were tested by Chi-square test and are statistically significant at $p < 0.001$ so omitted for clarity*

